# Osteophyte size and location on hip DXA scans are associated with hip pain: findings from a cross sectional study in UK Biobank

**DOI:** 10.1101/2021.04.26.21255905

**Authors:** Benjamin G. Faber, Raja Ebsim, Fiona R. Saunders, Monika Frysz, Claudia Lindner, Jennifer S. Gregory, Richard M. Aspden, Nicholas C. Harvey, George Davey Smith, Timothy Cootes, Jonathan H. Tobias

## Abstract

**Objective:** It remains unclear how the different features of radiographic hip osteoarthritis (rHOA) contribute to hip pain. We examined the relationship between rHOA, including its individual components, and hip pain using a novel dual-energy x-ray absorptiometry (DXA)-based method.

**Methods:** Hip DXAs were obtained from UK Biobank. An automated method was developed to obtain minimum joint space width (mJSW) from points placed around the femoral head and acetabulum. Osteophyte areas at the lateral acetabulum, superior and inferior femoral head were derived manually. Semi-quantitative measures of osteophytes and joint space narrowing (JSN) were combined to provide a measure of rHOA. Logistic regression was used to examine the relationships between these variables and hip pain, obtained via questionnaires.

**Results:** 6,807 hip DXAs were examined. rHOA was present in 353 [5.2%] individuals and was associated with hip pain [OR 2.07 (95% CI 1.54-2.80)] and hospital diagnosed OA [5.73 (2.89-11.36)]. Total osteophyte area and mJSW were associated with hip pain [1.29 (1.21-1.36), 0.84 (0.77-0.92) respectively] in unadjusted models. After mutually adjusting and adding demographic covariates, total osteophyte area continued to have strong evidence of association with hip pain [1.31 (1.23-1.39)] but mJSW did not [0.95 (0.87-1.04)]. Acetabular, superior and inferior femoral osteophyte areas were all independently associated with hip pain [1.19 (1.13-1.26), 1.22 (1.15-1.29), 1.21 (1.14-1.28) respectively].

**Conclusion:** The relationship between DXA-derived rHOA and prevalent hip pain is explained by osteophyte area rather than mJSW. Osteophytes at different locations showed important, potentially independent, associations with hip pain, possibly reflecting the contribution of distinct biomechanical pathways.

## Introduction

Osteoarthritis (OA) is a common condition with important sequelae in terms of morbidity and mortality, predominantly affecting knees, hands, spine and hip joints (1, 2). Hip OA (HOA) can be defined radiographically (rHOA) using classification systems such as Kellgren-Lawrence (KL) or Croft (3, 4). rHOA is comprised of joint space narrowing (JSN), osteophytes, subchondral sclerosis and cysts, of which JSN and osteophytes are most frequently recorded (3, 5, 6). rHOA is usually studied as a categorical variable (0-4 for KL scoring (3) or 0-5 Croft scoring (4)) with a threshold defined for the presence of rHOA. HOA can also be defined symptomatically (sHOA) (7, 8).

KL classification of rHOA has been shown to have a poor sensitivity and predictive value for symptoms (9). That said, severity of radiographic changes is associated with likelihood of symptoms and total hip replacement, a proxy for end-stage disease (10, 11). Previous studies have also examined the relationship between individual features of rHOA and hip pain, for example JSW was found to be only weakly associated with symptomatic measures of HOA (12). Another study examined the relationship between individual semi-quantitively graded components of rHOA and hip pain in women, observing that femoral head osteophytes were related to hip pain more strongly than JSN (10). A recent small study found that inferior medial femoral head osteophytes seen on computed tomography (CT) scans were associated with hip pain more strongly than other (superolateral, intra-articular, anterior and posterior) osteophytes, indicating that the relationship between osteophytes and hip pain may differ according to osteophyte location (13). With improving technology, it is now possible to measure features of rHOA in greater detail, for example measuring osteophyte size quantitively although this has not previously been applied to large population-based studies (14-16). By studying individual features of rHOA in greater detail this may help to better understand their contribution to the development of hip pain, providing a basis for more accurate diagnostic/prognostic imaging biomarkers, and greater understanding of the biomechanical pathways underpinning OA development.

To date, large epidemiological studies of rHOA have almost exclusively been based on radiographs using well recognised atlases (17). In contrast, dual-energy X-ray absorptiometry (DXA) hip scans, widely used to evaluate patients for osteoporosis, and obtained in many large cohort studies, have previously had insufficient resolution to evaluate features related to osteoarthritis such as osteophytes (6). However, a new generation of DXA machines is now available with resolution comparable with that of radiographs, which have been validated for KL grading (18). This opens up the possibility of using cohort studies, in which large numbers of individuals have undergone newer generation hip DXA scans, to study rHOA; such as the UK Biobank (UKB) extended imaging study due to comprise 100,000 individuals (19, 20). Here, we aimed to evaluate the feasibility of this approach, by deriving a measure of rHOA in a subset of 7000 hip DXA scans from UKB and, relating this to previously diagnosed HOA and hip pain. Further, we examined the relationship between hip pain and the different elements of rHOA in this substantial sample, and hip pain, including the contribution of osteophyte size and location.

## Materials and Methods

### Population

UKB is a prospective mixed sex cohort based in the UK which recruited 500,000 adults aged 40-69 years old between 2006-2010. All participants underwent extensive physical, health and genetic phenotyping through electronic questionnaires, physical measurements and bodily fluid analysis (21). UKB is overseen by the Ethics Advisory Committee and received approval from the National Information Governance Board for Health and Social Care and North West Multi-centre Research Ethics Committee (11/NW/0382), all participants provided informed consent for this study which was approved by UKB (application number 17295). A full data catalogue is available online (http://biobank.ctsu.ox.ac.uk/crystal/). In 2013, the extended imaging study started which aims to conduct hip and whole body DXA scans on 100,000 of the participants; to date over 45,000 individuals have been scanned (19). DXA scans of both hips (iDXA GE-Lunar, Madison, WI) were obtained from participants positioned with 15-25° internal rotation using a standardised protocol (22). This study is based on a random sub-sample of 7000 individuals, selected from the overall sample of 13,496 individuals with DXA scans available at the time (February 2020). The first 20% of the subsample were selected randomly from those with a self-reported diagnosis of OA (the question did not ask at which joints) with the aim of increasing the number of pathological scans for our automated model training as part of a wider research programme. The remainder of the sample (80%) was selected randomly, throughout randomisation was achieved using a random number generator whilst we ensured the sexes were split equally.

Across all UKB participants 8.6% have self-reported a diagnosis of OA. All demographic information was taken from questionnaires completed on the same day as the DXA scan. Ethnicity was self-reported, and individuals were categorised into white, Asian, black, mixed-heritage, Chinese and other. The participants were asked via electronic questionnaire; *“Have you had hip pains for more than 3 months?”* They could answer “yes”, “no”, “don’t know”, “prefer not to say” or leave the answer blank, for this study only those who answered “yes” were categorised to have hip pain and the rest were not. Of note the hip pain question was not side specific. Hospital episode statistics linked with UKB were reviewed for ICD-9 & -10 codes related to HOA and if any were present then the individual was categorised to have hospital diagnosed HOA, as a binary variable.

### DXA and osteophyte mark up

The left hip DXA was examined from each participant, 85 outline points were placed around the outline of the superior acetabulum, femoral head and metaphysis, lesser and greater trochanters by an automated Random Forest-based machine-learning algorithm before being reviewed and corrected where necessary by 4 manual annotators (23). 18 key points were anatomically guided, and the remaining points were equally spaced between these (Supplementary Figure S1).

A DXA-based atlas was created by BF, FS and MW (see acknowledgements) describing osteophytes at the lateral acetabulum, superolateral femoral head and inferomedial femoral head, based on the OARSI radiographic atlas (17). Femoral head osteophytes are referred to as superior and inferior femoral head osteophytes for simplicity. Two annotators (BF & FS) examined all the images to mark-up osteophytes, using a custom tool (The University of Manchester) to mark each osteophyte area and move the outline points inside of the osteophyte margin (Figure 1). All osteophytes and adjoining points were agreed between these two annotators. The area of each osteophyte in millimetres squared (mm^2^) was then derived for each image to be used as a continuous variable describing osteophyte size. The osteophytes from the first 1930 DXAs were semi-quantitatively graded (grade 1-3) based on the aforementioned DXA-based atlas. Receiver operating characteristic curves (ROC) were used to define a threshold using osteophyte area scores for grade ≥1 and grade ≥2 osteophytes at each location to automate semi-quantitative grading of the remaining images (the presence of a grade 1 osteophyte was set at a threshold of osteophyte area ≥1mm^2^ at all locations, area under the curve (AUC) 1; acetabular grade ≥2 osteophyte: threshold ≥10mm^2^, AUC 0.96; superior femoral grade ≥2 osteophyte: threshold ≥17mm^2^, AUC 0.98; inferior femoral grade ≥2 osteophyte: threshold ≥19mm^2^, AUC 1). It was necessary to combine manually graded 2 and 3 osteophytes due to low numbers of grade 3 osteophytes (grade 3 osteophytes by location: acetabular n = 11, superior femoral head n = 6, inferior femoral head n = 4).

**Figure 1:**
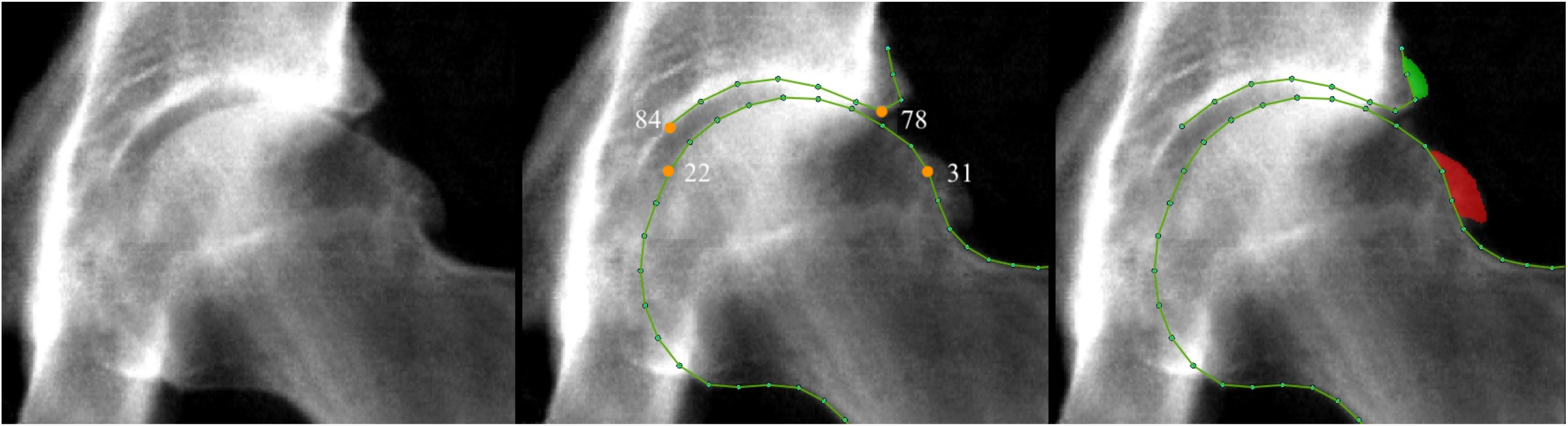
An example of a DXA image from UK Biobank. Left image: This is an example of a high-resolution hip DXA from UK Biobank showing radiographic osteoarthritis. Middle image: This shows how the points were placed on the borders of the bone on the same image. Points 22, 31, 78 and 84 are labelled and orange showing the area over which minimum joint space width was measured. Right image: This shows the acetabular osteophyte (green) and superior femoral head osteophyte (red) marked up on the same image.

### Joint Space Width

An automated method for measuring the width of the superior joint space, which is well demarcated on DXA (Figure 1), was subsequently developed. A custom Python script calculated mJSW between the acetabulum (points 78-84) and superior femoral head (points 22-31) as follows: A segment is created by drawing a straight line between two neighbouring points, for example, two points on the acetabulum. Then the shortest distance is calculated between this line and an opposing point, in this example on the femoral head. The automated method repeats this process for all segments and points selected, and the shortest distance representing mJSW (in mm) is saved. Additionally, the first 1930 DXAs were semi-quantitatively graded for JSN, blinded to mJSW, using a DXA-based JSN atlas created by BF, FS & MW, based on the OARSI atlas (17). Height-adjusted ROC curves were used to define thresholds for JSN automatically on the remaining images, as these thresholds were found to be more accurate at defining JSN than from mJSW alone, giving AUC 0.92 for JSN grade ≥1 and 0.97 for grade ≥2. Grades 2 & 3 were merged due to the low numbers of grade 3 JSN (n=9).

### Radiographic hip osteoarthritis

rHOA was defined as grade ≥1 JSN combined with a grade ≥1 osteophyte(s), as this was felt to be most equivalent to Kellgren-Lawrence and Croft definitions based on JSN combined with a definite osteophyte(s) (3, 4). Subchondral sclerosis and cysts were not examined as part of this study due to their relative infrequency (5). A more stringent definition of rHOA termed grade ≥2 rHOA, was defined as grade ≥2 osteophyte(s) combined with grade ≥ 2 JSN.

### Statistical analysis

The demographic data are given as a mean and range for continuous variables and binary variables are given as counts and frequency. The initial analyses investigated categorical measures of rHOA, osteophytes, JSN and hip pain using logistic regression with results presented as odds ratios (OR) with 95% confidence intervals (CI). Later analyses examined continuous measures of osteophyte area and mJSW against hip pain again using logistic regression. Use of directed acyclic graphs informed the *a priori* selection of covariates, which included age, sex, height, weight and ethnicity to be added into an adjusted model. Logistic regression was also used to examine the independent relationships between rHOA features and hip pain through mutually adjusted models. Graphical representations of logistic regression models were created by deriving the probability of hip pain from the regression model at specific intervals of osteophyte area or mJSW, and plotting these. We refer to this as the likelihood of hip pain rather than probability to avoid confusion with P-values. All statistical analysis was performed using Stata version 15 (StataCorp, College Station, TX, USA).

## Results

### Descriptives: Population characteristics

Of the initial sample of 7000 participants with a left hip DXA, 193 were excluded (72 had a significant artefact, 39 were missing the greater trochanter, 32 were missing the lesser trochanter, 29 were missing part of the femoral head or femur, 3 were missing part of the ilium or acetabulum, 16 were poor quality, and 2 individuals withdrew consent for the study). This left a total of 6,807 individuals (mean age 62.7 years old, standard deviation (SD) 7.5 years) with left hip DXAs available for analysis (Table 1). The sample was made up of 3425 [50.3%] females and 3382 [49.7%] males. 1489 [21.9%] self-reported a diagnosis of OA (no joint locations were specified in the question), 594 [8.7%] reported hip pain for more than 3 months at the time of imaging study attendance and 47 [0.7%] had hospital-diagnosed OA.

**Table 1:** Demographics of the sample studied with grade ≥1 abnormalities included. Abbreviations: Osteoarthritis (OA), radiographic hip osteoarthritis (rHOA), joint space narrowing (JSN), osteophyte (OP), joint space width (JSW).

|  | Males | Females | Combined |
| --- | --- | --- | --- |
| **Demographics** | Mean [Range] | Mean [Range] | Mean [Range] |
| Age (years) | 63.4 [45-80] | 62.1 [46-79] | 62.7 [45-80] |
| Weight (kg) | 83.8 [50-160] | 68.7 [36-155] | 76.2 [36-160] |
| Height (cm) | 177.0 [153-203] | 163.3 [137-195] | 170.1 [137 – 203] |
| Hip Pain | 219 [6.5] | 375 [11.0] | 594 [8.7] |
| Self-reported OA | 581 [17.2] | 908 [26.5] | 1489 [21.9] |
| *Ethnicity* | Prevalence [%] | Prevalence [%] | Prevalence [%] |
| White | 3278 [97.0] | 3321 [97.0] | 6599 [97.0] |
| Asian | 48 [1.4] | 26 [0.8] | 74 [1.1] |
| Black | 23 [0.7] | 20 [0.6] | 43 [0.6] |
| Mixed heritage | 13 [0.4] | 21 [0.6] | 34 [0.5] |
| Chinese | 5 [0.2] | 9 [0.3] | 14 [0.2] |
| Unknown | 15 [0.4] | 28 [0.8] | 43 [0.6] |
| *rHOA measures* | Prevalence [%] | Prevalence [%] | Prevalence [%] |
| rHOA | 245 [7.2] | 108 [3.2] | 353 [5.2] |
| JSN | 817 [24.2] | 543 [15.9] | 1360 [20] |
| Any OP | 709 [21.0] | 448 [13.1] | 1157 [17] |
| Acetabular OP | 484 [14.3] | 345 [10.1] | 829 [12.2] |
| Superior Femoral OP | 289 [8.6] | 143 [4.2] | 432 [6.4] |
| Inferior Femoral OP | 168 [5.0] | 52 [1.5] | 220 [3.2] |
| OP All | 45 [1.3] | 16 [0.5] | 61 [0.9] |
| Minimum JSW (mean [range]) | 2.9 [0.3 – 5.9] | 2.7 [0.2 – 4.8] | 2.8 [0.2 – 5.9] |
| **Total Sample** | 3382 | 3425 | 6807 |

### Descriptives: Features of rHOA

Prevalent rHOA, defined as grade ≥1 osteophyte combined with grade ≥1 JSN, was present in more males [245 (7.2%)] than females [108 (3.2%)] (Table 1). Mean mJSW, defined as the narrowest point of superior joint space, was 2.9 mm (SD 0.6 mm) and 2.7 mm (SD 0.5 mm) in males and females respectively. Grade ≥1 JSN was more common in males [817 (24.2%)] than females [543 (15.9%)]. Grade ≥1 osteophytes were recorded in 1157 [17%] individuals with the most common site being the lateral acetabulum [829 (12.2%)], followed by the superior femoral head [432 (6.4%)] and inferior femoral head [220 (3.2%)] with 61 [0.9%] individuals having an osteophyte at all three sites. Osteophytes were more frequently seen in males [709 (21%)] than females [448 (13.1%)] (Table 1). Supplementary Table S1 shows comparable descriptions for grade ≥2 rHOA defined by grade ≥2 osteophytes combined with grade ≥2 JSN. In terms of continuous measures of osteophytes in those individuals with osteophytes, mean total area of all osteophytes present was 25 mm^2^ with a range from 2 mm^2^ to 268 mm^2^. Mean area of individual osteophytes was 16 mm^2^ (range 2-157 mm^2^), 24 mm^2^ (3-121 mm^2^) and 21 mm^2^ (2-157 mm^2^) for lateral acetabular, superior femoral head and inferior femoral head osteophytes respectively.

### rHOA versus self-reported OA and hip pain

In unadjusted analyses, rHOA and grade ≥2 rHOA were associated with self-reported diagnosis OA [OR 1.53 (95% CI 1.21-1.94) and 1.97 (1.36-2.84) respectively]. These associations strengthened slightly after adjustment for demographic covariates, namely age, sex, height, weight and ethnicity [OR 1.68 (1.31-2.15) and 2.12 (1.45-3.10) respectively]. In unadjusted analyses, rHOA and grade ≥2 rHOA were also associated with a hospital diagnosis of HOA [OR 5.73 (2.89-11.36) and 7.96 (3.32-19.10) respectively], with similar results after adjustment for demographic covariates [OR 6.01 (2.98–12.16) and 9.02 (3.60-22.62) respectively]. In unadjusted analyses, rHOA was associated with prevalent hip pain [OR 2.07 (1.54-2.80)], with similar results after adjustment for demographic covariates (Table 2). Stronger associations were observed between grade ≥2 rHOA and hip pain [OR 3.17 (2.08-4.84)] (Supplementary Table S2).

**Table 2:** The associations between radiographic hip osteoarthritis and its constituent features, and hip pain. Logistic regression comparing the presence of radiographic hip osteoarthritis (rHOA) and its constituent features and hip pain in 6807 individuals. Odd ratios (OR) presented with 95% confidence intervals (CI) and P-values. rHOA defined as the presence of grade ≥1 joint space narrowing (JSN) and a grade ≥1 osteophyte (OP). Any OP refers to a grade ≥1 OP at any site (binary measure). OP presence at each location is examined as Acetabular OP, Superior Femoral OP, Inferior Femoral OP. OP at all 3 sites refers to concurrent OPs at all sites examined. Hip pain (yes/no) derived from questionnaire data taken on the same day as DXA scan. Unadjusted and adjusted results shown. Adjusted model includes age, sex, height, weight, ethnicity.

|  | Hip Pain |  |  |  |
| --- | --- | --- | --- | --- |
|  | Unadjusted |  | Adjusted |  |
|  | OR [95% CI] | *P* | OR [95% CI] | *P* |
| rHOA | 2.07 [1.54-2.8] | $1.74 \times 10^{-06}$ | 2.42 [1.78-3.29] | $1.59 \times 10^{-08}$ |
| JSN | 1.18 [0.97-1.45] | 0.10 | 1.30 [1.06-1.60] | 0.01 |
| Any OP | 1.64 [1.35-2.01] | $1.06 \times 10^{-06}$ | 1.73 [1.41-2.13] | $1.20 \times 10^{-07}$ |
| Acetabular OP | 1.67 [1.33-2.09] | $6.50 \times 10^{-06}$ | 1.69 [1.35-2.12] | $6.06 \times 10^{-06}$ |
| Superior Femoral OP | 2.20 [1.68-2.88] | $9.90 \times 10^{-09}$ | 2.51 [1.91-3.31] | $6.17 \times 10^{-11}$ |
| Inferior Femoral OP | 2.58 [1.82-3.65] | $8.91 \times 10^{-08}$ | 3.09 [2.16-4.42] | $6.44 \times 10^{-10}$ |
| OP at all 3 sites | 6.09 [3.60-10.34] | $2.30 \times 10^{-11}$ | 7.14 [4.15-12.30] | $1.30 \times 10^{-12}$ |

### Osteophytes and joint space width (CATEGORICAL measures) versus hip pain

The presence of a grade ≥1 osteophyte at any site was associated with hip pain [OR 1.64 (1.35-2.01)] in unadjusted analyses, which were unaffected by adjustment as above (Table 2). Grade ≥2 osteophytes at any location demonstrated a greater relationship with hip pain [OR 1.99 (1.57-2.52)] (Supplementary Table S2). Unadjusted analyses showed no evidence of association between grade ≥1 JSN and hip pain (Table 2). However, grade ≥2 JSN was associated with hip pain, in both unadjusted and adjusted analyses (Supplementary Table S2). In unadjusted analyses, the presence of grade ≥1 acetabular osteophytes [OR 1.67 (1.33-2.09)], superior femoral osteophytes [OR 2.20 (1.68-2.88)] and inferior femoral osteophytes [OR 2.58 (1.82-3.65)] were all associated with prevalent hip pain and this did not alter with adjustment for demographic covariates (Table 2). The relationships for each osteophyte site were only minimally attenuated by additional mutual adjustment [acetabular osteophyte OR 1.40 (1.10-1.78), superior femoral osteophyte OR 1.86 (1.36-2.54), inferior femoral osteophyte OR 2.01 (1.35-3.00)]. Individuals with osteophytes at all three sites showed stronger associations with hip pain in both unadjusted [OR 6.09 (3.60-10.34)] and adjusted analyses (Table 2). Grade ≥2 osteophytes had a greater association with prevalent hip pain [acetabular osteophyte OR 2.08 (1.59-2.72), superior femoral osteophyte OR 2.62 (1.90-3.62), inferior femoral osteophyte OR 5.53 (3.39-9.02), all 3 osteophytes OR 14.97 (6.62-33.86) (unadjusted analyses)] (Supplementary Table S2). Sex-stratified results showed similar associations between features of rHOA and hip pain in males and females (Supplementary Tables S3 & S4).

### Osteophytes and joint space width (CONTINUOUS measures) versus hip pain

Total osteophyte area was associated with prevalent hip pain in unadjusted analyses [OR 1.29 (per standard deviation (SD) increase in area) (1.21-1.36)] (Figure 2). mJSW was also associated with hip pain in unadjusted analyses [OR 0.84 (per SD increase in width) (0.77-0.92)], the negative association conferring an increased risk of pain with decreasing JSW. To examine independent effects total osteophyte area and superior mJSW were combined in a mutually adjusted single model. Total osteophyte area remained strongly associated with hip pain [OR 1.27 (1.19-1.34)], but the association with superior mJSW was marginally attenuated [OR 0.90 (0.83-0.98)] (Supplementary Figure S2). The addition of demographic covariates had little effect on the association between total osteophyte area and hip pain [OR 1.31 (1.23-1.39)] but attenuated the association with superior mJSW and hip pain towards the null [OR 0.95 (0.87-1.04)] (Figure 2).

**Figure 2:**
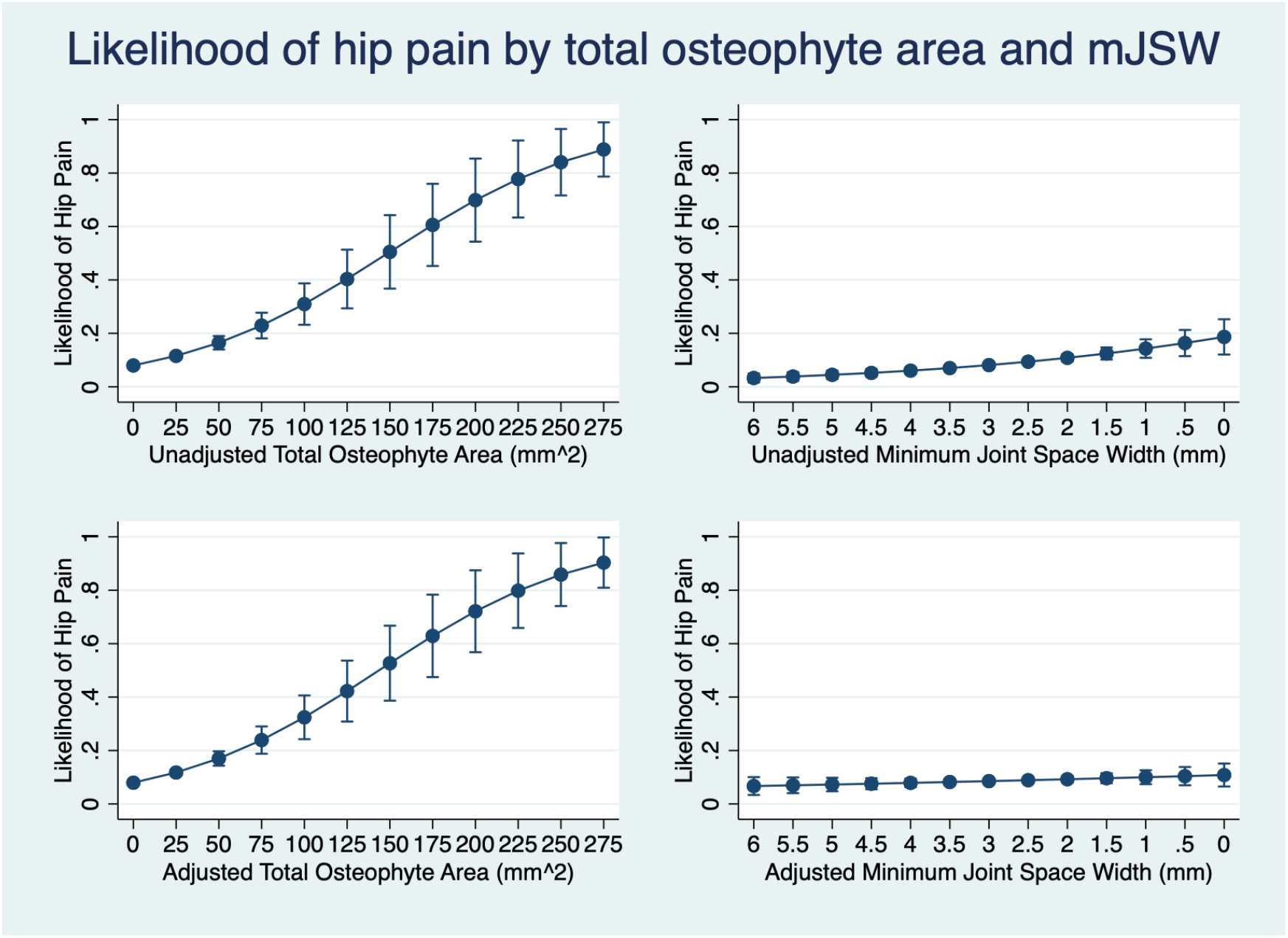
Likelihood of hip pain depending on total osteophyte area and minimum joint space width. Top left graph shows the unadjusted likelihood of hip pain by total osteophyte area. Top right graph shows the unadjusted likelihood of hip pain by mJSW, the x-axis is reversed. Bottom left graph shows likelihood of hip pain by total osteophyte area, adjusted for mJSW, age, sex, height, weight and ethnicity. Bottom right graph shows likelihood of hip pain by mJSW, adjusted for total osteophyte area, age, sex, height, weight and ethnicity.

Osteophyte area at specific sites was associated with hip pain [acetabular osteophyte area OR 1.19 (per SD increase) (1.13-1.26), superior femoral osteophyte area OR 1.22 (1.15-1.29), inferior femoral osteophyte area OR 1.21 (1.14-1.28) (unadjusted analyses)] (Figure 3). When regional osteophyte areas were mutually adjusted for each other in a combined model, acetabular osteophyte area [OR 1.13 (1.06-1.20)], superior femoral osteophyte area [OR 1.13 (1.05-1.24)] and inferior femoral osteophyte area [OR 1.10 (1.03-1.17)] remained associated with hip pain (Supplementary Figure S3). Similar results were observed following additional adjustment for demographic covariates [acetabular osteophyte area OR 1.13 (1.06-1.21), superior femoral osteophyte area OR 1.16 (1.08-1.24) and inferior femoral osteophyte area OR 1.11 (1.04-1.19)] (Figure 3).

**Figure 3:**
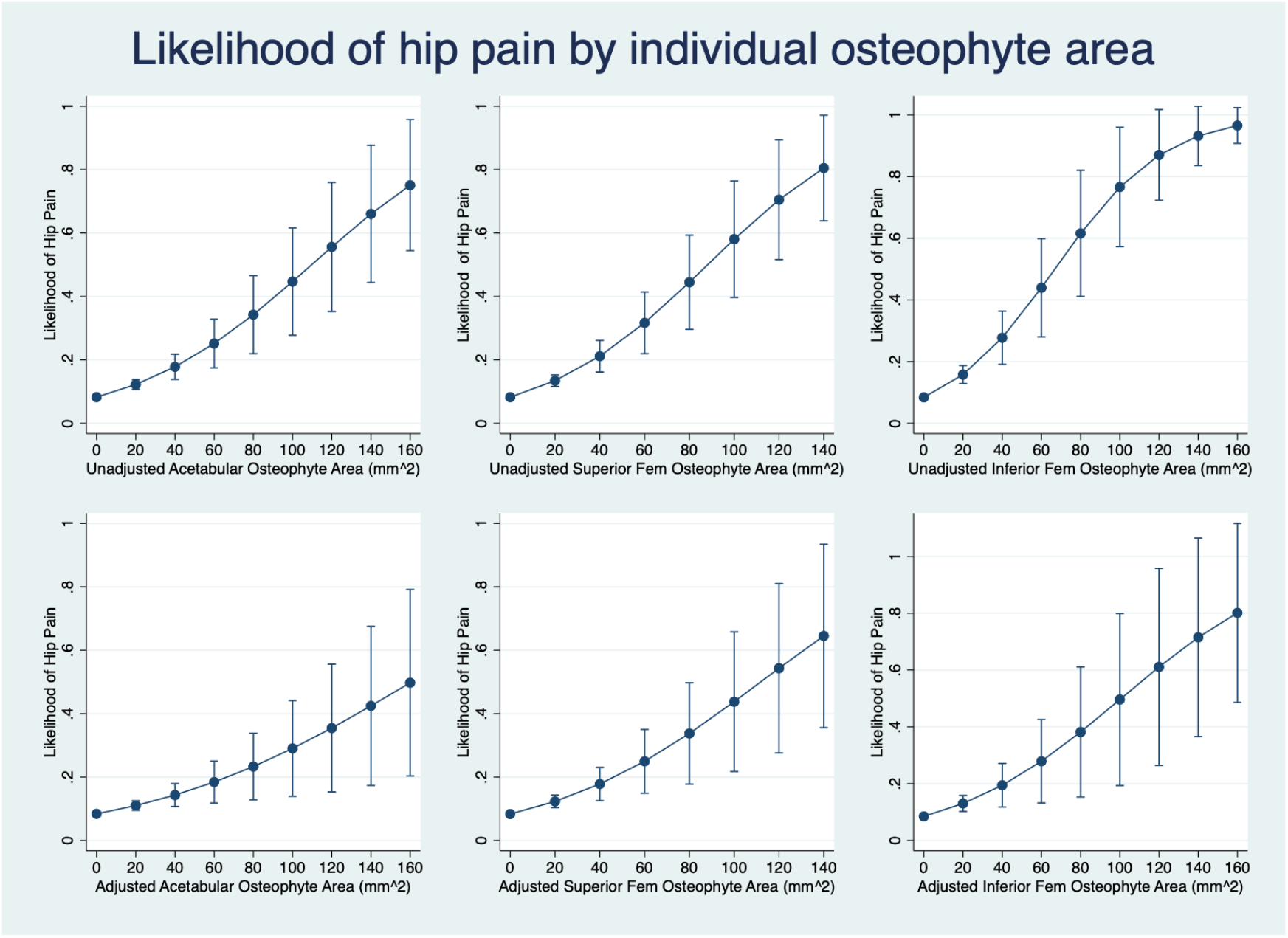
Likelihood of hip pain depending on regional osteophyte area. Top left graph shows the unadjusted likelihood of hip pain by acetabular osteophyte area. Top middle graph shows the unadjusted likelihood of hip pain by superior femoral osteophyte area. Top right graph shows the unadjusted likelihood of hip pain by inferior femoral osteophyte area. The corresponding graphs below represent the respective models adjusted for area of osteophytes at the other sites, age, sex, height, weight and ethnicity.

## Discussion

In a large (n = 6,807) cross-sectional study of both men and women, we have developed and applied a method for performing detailed phenotyping of rHOA based on high resolution DXA scans. As expected, those with rHOA as defined by DXA were associated with a higher prevalence of self-reported and hospital-diagnosed OA. We then went on to explore the relationship between rHOA and its individual features, and prevalent hip pain. We found that DXA-derived rHOA is associated with prevalent hip pain and that this association is predominately driven by the presence of osteophytes, rather than joint space narrowing. Subsequently, we examined the relationship between osteophytes and hip pain based on quantitative evaluations of osteophyte size and osteophyte location. We found a positive relationship between osteophyte area and the likelihood of hip pain, such that the latter exceeded 50% when total osteophyte area reached 150 mm^2^, implying florid osteophytes are most reliably associated with hip pain. In addition, we found that osteophytes at all three sites examined, namely acetabular, superior femoral and inferior femoral, all showed potentially independent relationships with hip pain, consistent with roles in partially-independent biomechanical pathways. Inferior femoral osteophytes showed the strongest association with hip pain, and acetabular osteophytes the weakest.

Previous studies have shown that rHOA is poorly predictive of hip pain but these have focused on semi-quantitative composite measures of rHOA which may have limited accuracy in the assessment of joint pathology (9). Semi-quantitative measures of rHOA generally group together different osteophyte locations and sizes and use broad definitions of JSN, which may partly explain the weak associations observed with symptoms at both hip and knee joints (9, 24-26). We observed similar findings in our analysis, as even though individuals who had either DXA-derived rHOA or a single osteophyte (grade ≥1) were at an elevated risk of hip pain, it was still the case that the majority of them did not have any hip pain (84% and 88% respectively). We are not aware of any previous studies to have examined clinical outcomes in relation to quantitative measures of hip osteophyte size as presented here. However there have been two previous studies analysing the relationship between osteophyte location and hip pain, with which our results are consistent. One previous study (n = 5,839) found that femoral osteophytes have a greater association with hip pain compared to acetabular osteophytes in women (10). A small CT-based study (n = 29) found that inferior osteophytes had a stronger association with hip pain compared with anterior, posterior and intra-articular osteophytes (13).

Osteophytes are a key component of OA although little is known about if or how they might induce pain, with many patients who have osteophytes not suffering from pain (27). Kijima et al. suggest that inferior femoral head osteophytes are a proxy for hip instability which might be causing hip pain through impingement of the femoral head and acetabulum (13). It is known that osteophytes are a poor prognostic sign for arthroscopic interventions for hip pain potentially due to a stabilising effect they have on a joint which is lost if they are removed (24, 28). Others have shown that osteophytes contain sensory fibres suggesting pain could be derived from the osteophyte itself (29, 30), although arthroscopic removal of osteophytes is ineffective in the treatment of knee pain and no longer recommended (31, 32). In addition, pain might be associated with osteophytes due to periostitis or inflammation which leads to their development rather than the osteophyte itself causing pain (33).

Our analysis, showing independent relationships between osteophytes at different sites and hip pain suggests location-specific mediators are a possibility, such as a role of distinct biomechanical pathways. Along similar lines, associations between hip morphology and rHOA and risk of hip replacement are presumed to be mediated through aberrant biomechanical pathways (6, 34, 35). How such variations in morphology are related to specific constituents of rHOA remains unclear. Studies from high bone mass individuals show a global predisposition to osteophyte formation (hypertrophic OA), suggesting a strong genetic influence on osteophyte formation (5, 36), which might point against specific local biomechanical factors in the development of osteophytes. On the other hand, it could still be the case that osteophytes lead to pain through local mechanisms as suggested by the independent relationships seen in this study. Understanding if and how different osteophytes contribute to pain is of clear clinical interest and requires further investigation.

Superior mJSW was associated with hip pain in our unadjusted model, but the relationship attenuated after adjustment for total osteophyte area and demographic covariates. These findings are consistent with a previous systematic review which only found weak associations between JSW and hip pain (12). A previous study on incident knee OA in a high bone mass population found that change in Western Ontario and McMaster Universities Osteoarthritis Index (WOMAC) pain score over time was attenuated to a greater extent by adjustment for osteophyte score, compared with joint space narrowing (36), further suggesting that osteophytes are the main contributing factor to the relationship between rHOA and joint pain. To the extent that JSW contributes a limited amount to the evolution of hip pain in rHOA, this would seemingly undermine its use as an endpoint in clinical trials of disease modifying osteoarthritis drugs (DMOAD) (37).

A major strength of this study was the use of a novel method for characterising different components of rHOA on DXA scans, developed as part of our investigation This enabled us to examine relationships between detailed measures of rHOA and hip pain in a large sample of participants from UKB. Although there are limited data available on the validity of using hip DXA scans to ascertain rHOA, the measures we obtained showed expected relationships with hospital-diagnosed and self-reported OA. Whilst DXA scan images appear suitable for deriving characteristics such as osteophytes and superior joint space width, including the potential for automation, they have several inherent limitations in evaluating rHOA. A potential limitation in the use of DXA scans to measure joint space width is that scans are obtained with the patient supine, rather than weight bearing as is the norm for radiographs (38). However, a previous study found little difference in JSW between weight bearing and non-weight bearing hip radiographs (39). Limitations in DXA imaging prevented us from evaluating other radiographic features associated with rHOA, such as subchondral sclerosis and cysts which were difficult to visualise. In addition, in contrast to the superior joint space, we were unable to visualise or evaluate the medial or inferior joint space as is often possible on x-rays.

The limitations of this study include, the observational and cross-sectional study-design which makes it not a suitable basis for drawing causal conclusions. In particular we can only comment on relationships with prevalent rather than incident hip pain. The hip pain information is limited in that it is not side-specific, although it does cover a prolonged duration (≥3 months) which makes it pertinent to HOA (33). Further, this study used a weighted sample to include a greater proportion of individuals with self-reported OA which means we cannot use this data to comment on the prevalence of rHOA in UKB.

To conclude, we have developed and applied a method for large scale phenotyping of rHOA on DXA scans in UKB. The measures of rHOA obtained showed expected relationships with clinical outcomes such as hip pain. Focusing on individual semi-quantitatively graded features, JSN and osteophytes at different sites, these showed associations with hip pain. On examining these relationships in more detail, based on quantitative measures derived for osteophyte area and mJSW, we found that mJSW had no independent association with hip pain, in contrast to osteophytes which showed potentially independent relationships at all three sites. Further studies are justified to characterise site-specific biomechanical alterations that result in or from the formation of osteophytes, to further understand if and how these changes might be causally related to symptoms of pain in HOA, and to explore whether their correction might provide a means of slowing OA progression.

## Data Availability

The data from this study will be available from UK Biobank at a forthcoming data release. Users must be registered with UK Biobank to access their resources [https://bbams.ndph.ox.ac.uk/ams/].

## Acknowledgements

The authors would like to thank Dr Martin Williams, Consultant Musculoskeletal Radiologist North Bristol NHS Trust, who provided substantial training and expertise for this study. This work has been conducted using the UK Biobank resource, access application 17295.

