## Supplementary data for "Osteophyte size and location on hip DXA scans are associated with hip pain: findings from a cross sectional study in UK Biobank"

**Supplementary Material**

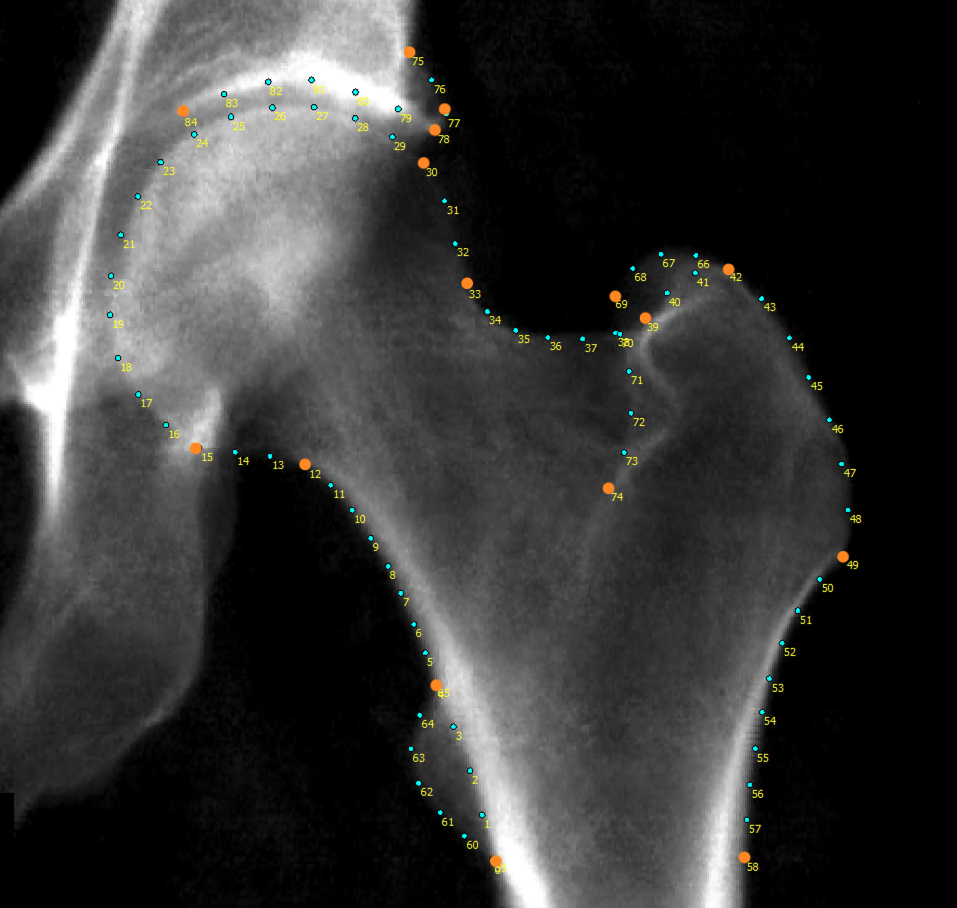
Supplementary Figure S1: A UKB hip DXA with numbered points placed around the joint and key points are highlighted in orange. Points 4&65 and 0&59 overlap in this example.

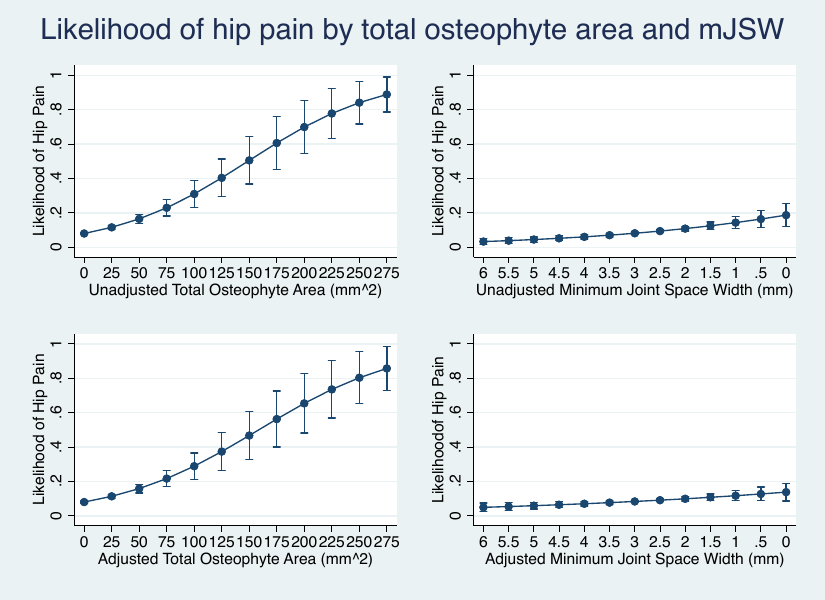
Supplementary Figure S2: Likelihood of hip pain depending on total osteophyte area and minimum joint space width (mJSW). Top left graph shows the unadjusted likelihood of hip pain by total osteophyte area. Top right graph shows the unadjusted likelihood of hip pain by mJSW, the x-axis is reversed. Bottom left graph shows the adjusted likelihood of hip pain by total osteophyte area adjusted for mJSW only. Bottom right graph shows the adjusted likelihood of hip pain by mJSW adjusted for total osteophyte area only.

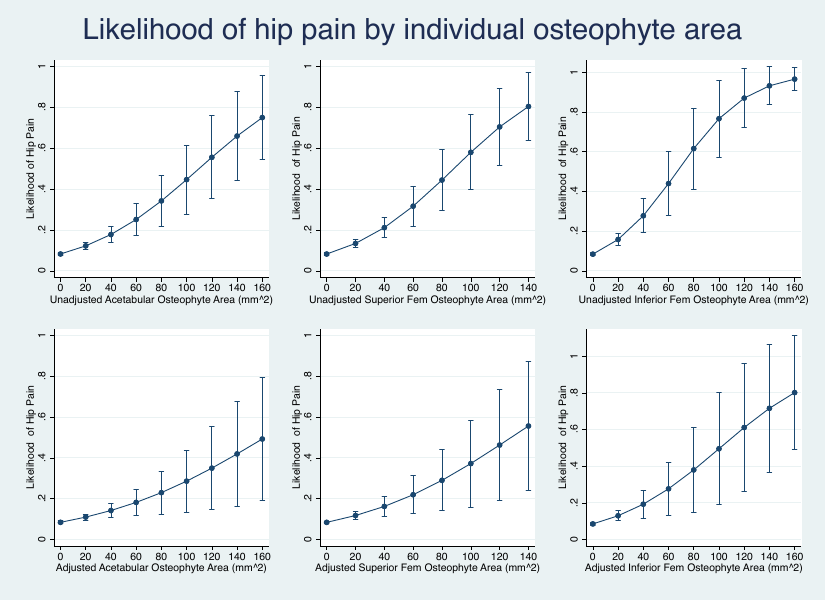
Supplementary Figure S3: Likelihood of hip pain depending on regional osteophyte area. Top left graph shows the unadjusted likelihood of hip pain by acetabular osteophyte area (mean 16.2 mm^2^). Top middle graph shows the unadjusted likelihood of hip pain by superior femoral osteophyte area (mean 23.8 mm^2^). Top right graph shows the unadjusted likelihood of hip pain by inferior femoral osteophyte area (mean 20.8 mm^2^). The corresponding graphs below represent the respective adjusted models, including the additional osteophyte areas only.

Supplementary Table S1. Demographics for sample based on grade ≥2 radiographic hip osteoarthritis.

|  | Males | Females | Combined |
| --- | --- | --- | --- |
| *rHOA binary measures* | Prevalence [%] | Prevalence [%] | Prevalence [%] |
| rHOA | 105 [3.1] | 23 [0.7] | 128 [1.9] |
| JSN | 338 [10.0] | 138 [4.0] | 476 [7.0] |
| Any OP | 431 [12.7] | 214 [6.5] | 645 [9.5] |
| Acetabular OP | 294 [8.7] | 164 [4.8] | 458 [6.7] |
| Superior Femoral OP | 177 [5.2] | 78 [2.3] | 255 [3.8] |
| Inferior Femoral OP | 53 [1.6] | 21 [0.6] | 74 [1.1] |
| OP All | 17 [0.5] | 7 [0.2] | 24 [0.4] |
| **Total Sample** | 3382 | 3425 | 6807 |

Supplementary Table S2. Logistic regression comparing the presence of grade ≥2 radiographic hip osteoarthritis (rHOA) and its constituent features and hip pain in 6807 individuals. Odd ratios (OR) presented with 95% confidence intervals (CI) and P-values. Grade ≥2 rHOA defined as the presence of grade ≥2 joint space narrowing (JSN) and a grade ≥2 osteophyte (OP). Any OP refers to a grade ≥2 OP at any site (binary measure). Grade ≥2 OP presence at each location is examined as Acetabular OP, Superior Femoral OP, Inferior Femoral OP. OP at all 3 sites refers to concurrent grade ≥2 OPs at all sites examined. Hip pain (yes/no) derived some questionnaire data taken on the same day as DXA scan. Unadjusted and adjusted results shown. Adjusted model includes age, sex, height, weight, ethnicity.

|  | Hip Pain | | | |
| --- | --- | --- | --- | --- |
|  | Unadjusted | | Adjusted | |
|  | OR [95% CI] | *P* | OR [95% CI] | *P* |
| rHOA | 3.17 [2.08-4.84] | 8.84 x 10^-08^ | 3.85 [2.49-5.95] | 1.33 x 10^-09^ |
| JSN | 1.53 [1.15-2.04] | 3.50 x 10^-03^ | 1.8 [1.34-2.42] | 9.27 x 10^-05^ |
| Any OP | 1.99 [1.57-2.52] | 1.03 x 10^-08^ | 2.17 [1.70-2.76] | 3.98 x 10^-10^ |
| Acetabular OP | 2.08 [1.59-2.72] | 7.35 x 10^-08^ | 2.16 [1.65-2.84] | 3.19 x 10^-08^ |
| Superior Femoral OP | 2.62 [1.90-3.62] | 5.31 x 10^-09^ | 3.05 [2.19-4.25] | 4.72 x 10^-11^ |
| Inferior Femoral OP | 5.53 [3.39-9.02] | 7.49 x 10^-12^ | 6.14 [3.72-10.16] | 1.50 x 10^-12^ |
| OP at all 3 sites | 14.97 [6.62-33.86] | 8.00 x 10^-11^ | 17.30 [7.53-39.74] | 1.90 x 10^-11^ |

|  | Males | | | | Females | | | |
| --- | --- | --- | --- | --- | --- | --- | --- | --- |
|  | Unadjusted | | Adjusted | | Unadjusted | | Adjusted | |
|  | OR [95% CI] | *P* | OR [95% CI] | *P* | OR [95% CI] | *P* | OR [95% CI] | *P* |
| rHOA | 2.78 [1.89-4.08] | 1.78 x 10^-07^ | 2.84 [1.92-4.18] | 1.52 x 10^-07^ | 1.90 [1.15-3.12] | 0.01 | 1.97 [1.19-3.26] | 8.16 x 10^-03^ |
| JSN | 1.45 [1.08-1.95] | 0.01 | 1.46 [1.08-1.97] | 0.01 | 1.15 [0.87-1.53] | 0.33 | 1.21 [0.91-1.61] | 0.20 |
| Any OP | 1.77 [1.31-2.39] | 1.78 x 10^-04^ | 1.73 [1.28-2.33] | 3.94 x 10^-04^ | 1.83 [1.39-2.41] | 1.56 x 10^-05^ | 1.78 [1.35-2.35] | 4.58 x 10^-05^ |
| Acetabular OP | 1.86 [1.34-2.59] | 2.40 x 10^-04^ | 1.80 [1.29-2.51] | 5.99 x 10^-04^ | 1.72 [1.26-2.34] | 5.41 x 10^-04^ | 1.65 [1.21-2.25] | 1.59 x 10^-03^ |
| Superior Femoral OP | 2.43 [1.68-3.54] | 2.90 x 10^-06^ | 2.56 [1.75-3.73] | 1.07 x 10^-06^ | 2.58 [1.72-3.87] | 4.50 x 10^-06^ | 2.59 [1.72-3.90] | 4.94 x 10^-06^ |
| Inferior Femoral OP | 3.01 [1.95-4.66] | 7.41 x 10^-07^ | 2.94 [1.89-4.58] | 1.72 x 10^-06^ | 3.39 [1.84-6.24] | 8.61 x 10^-05^ | 3.38 [1.82-6.26] | 1.10 x 10^-04^ |

Supplementary Table S3. Logistic regression comparing the presence of radiographic hip osteoarthritis (rHOA) and its constituent features and hip pain in 3382 males and 3425 females. Odd ratios (OR) presented with 95% confidence intervals (CI) and P-values. rHOA defined as the presence of grade ≥1 joint space narrowing (JSN) and a grade ≥1 osteophyte (OP). Any OP refers to an osteophyte at any site (binary measure). OP presence at each location is examined as Acetabular OP, Superior Femoral OP, Inferior Femoral OP. Hip pain (yes/no) derived some questionnaire data taken on the same day as DXA scan. Unadjusted and adjusted results shown. Adjusted model includes age, height, weight, ethnicity.

Supplementary Table S4 Logistic regression comparing the presence of grade ≥2 radiographic hip osteoarthritis (rHOA) and its constituent features and hip pain in 3382 males and 3425 females. Odd ratios (OR) presented with 95% confidence intervals (CI) and P-values. rHOA defined as the presence of grade ≥2 joint space narrowing (JSN) and a grade ≥2 osteophyte (OP). Any OP refers to a grade ≥2 osteophyte at any site (binary measure). Grade ≥2 OP presence at each location is examined as Acetabular OP, Superior Femoral OP, Inferior Femoral OP. Hip pain (yes/no) derived some questionnaire data taken on the same day as DXA scan. Unadjusted and adjusted results shown. Adjusted model includes age, height, weight, ethnicity.

|  | Males | | | | Females | | | |
| --- | --- | --- | --- | --- | --- | --- | --- | --- |
|  | Unadjusted | | Adjusted | | Unadjusted | | Adjusted | |
|  | OR [95% CI] | *P* | OR [95% CI] | *P* | OR [95% CI] | *P* | OR [95% CI] | *P* |
| rHOA | 4.14 [2.53-6.78] | 1.44 x 10^-08^ | 3.93 [2.39-6.49] | 8.01 x 10^-08^ | 3.61 [1.47-8.83] | 4.95 x 10^-03^ | 3.59 [1.46-8.84] | 5.49 x 10^-03^ |
| JSN | 1.86 [1.28-2.72] | 1.19 x 10^-03^ | 1.82 [1.24-2.66] | 2.14 x 10^-03^ | 1.67 [1.05-2.64] | 0.03 | 1.81 [1.13-2.89] | 0.01 |
| Any OP | 2.29 [1.65-3.18] | 8.42 x 10^-07^ | 2.26 [1.62-3.15] | 1.62 x 10^-06^ | 2.25 [1.59-3.20] | 5.63 x 10^-06^ | 2.14 [1.50-3.06] | 2.66 x 10^-05^ |
| Acetabular OP | 2.30 [1.58-3.35] | 1.42 x 10^-05^ | 2.2 [1.50-3.21] | 4.69 x 10^-05^ | 2.33 [1.58-3.44] | 2.12 x 10^-05^ | 2.18 [1.47-3.24] | 1.16 x 10^-04^ |
| Superior Femoral OP | 3.11 [2.03-4.75] | 1.59 x 10^-07^ | 3.34 [2.17-5.14] | 3.89 x 10^-08^ | 2.91 [1.73-4.89] | 5.79 x 10^-05^ | 2.93 [1.73-4.95] | 6.51 x 10^-05^ |
| Inferior Femoral OP | 6.66 [3.64-12.17] | 7.33 x 10^-10^ | 6.39 [3.46-11.79] | 2.92 x 10^-09^ | 6.23 [2.61-14.87] | 3.88 x 10^-05^ | 5.83 [2.41-14.08] | 9.00 x 10^-05^ |
